# Serum SARS-COV-2 Nucleocapsid Protein: A Sensitivity and Specificity Early Diagnostic Marker for SARS-COV-2 Infection

**DOI:** 10.1101/2020.05.24.20111849

**Authors:** Tao Li, Li Wang, Huihui Wang, Xuemei Li, Shubing Zhang, Yuanhong Xu, Wei Wei

## Abstract

**Objective:** This study aimed to explore the diagnostic value of serum severe acute respiratory syndrome coronavirus 2 (SARS-CoV-2) nucleocapsid(N) protein assay in the early stage of SARS-COV-2 infection.

**Method:** Serum N protein in SARS-COV-2 infected patients and non-SARS-COV-2 infected population was measured by enzyme-linked immunosorbent assay (ELISA) double antibody sandwich assay. Colloidal gold immunochromatography assay is used to detect serum N protein antibodies in the above population.

**Results:** 50 cases of SARS-CoV-2 nucleic acid positive and SARS-CoV-2 antibody negative patients had a serum N protein positive rate of 76%, including 2% with a concentration of 10.00-49.99 pg / mL, 8% with a concentration of 50.00-99.99 pg / mL, 22% with a concentration of 100.00 - 299.99 pg/mL, and 44% with a concentration≥ 300.00 pg / mL. 37 samples of patients with serum SARS-CoV-2 antibody positive after infection had a serum SARS-CoV-2 N protein positive rate of 2.7%, of which 2.7% had the concentration of 10.00-49.99 pg / mL and 0% had the concentration of 50.00-99.99 pg / mL, 100.00 −299.99 pg / mL, and ≥300.00 pg / mL. Serum N protein test results of 633 non-SARS-COV-2 infected patients including pregnant women, other respiratory infections, and increased rheumatoid factor were all negative, having a serum N protein concentration less than 10.00 pg/mL, with a specificity of 100%. Using SPSS 19.0 to calculate the receiver operating characteristic curve, the area under the curve was 0.9756 (95% confidence interval 0.9485-1.000, p <0.0001), sensitivity and specificity were 92% (95% confidence interval 81.16% to 96.85%) and 96.84% (95% confidence interval 95.17% to 97.15%). The best CUTOFF value is 1.850 pg / mL.

**Conclusion:** The measurement of SARS-COV-2 serum N protein has a high diagnostic value for the infected patients before the antibody appears, and shortens the window period of serological diagnosis. The laboratory needs to establish an individual CUTOFF value according to purpose of the application.

## Introduction

As of May 16, 2020, there were more than 5.1 million severe acute respiratory syndrome coronavirus 2 (SARS-CoV-2) infections and more than 333,000 deaths [1], which posed a serious threat to the health and economic life of people around the world. With the joint efforts of scientists all over the world, a variety of diagnostic reagents have been developed to provide solid support for clinical diagnosis [2-5]. At present, the diagnosis of SARS-COV-2 infection is mainly based on pharyngeal swab or sputum nucleic acid detection, and specific serum antibody detection is used as an auxiliary marker [6,7]. Nucleic acid testing is greatly affected by specimen collection, transportation and other stages. There are times of inconsistent positive and negative results, and the overall positive rate is not high [8,9], which brings great confusion to the clinical diagnosis. The detection of specific antibodies against SARS-COV-2 in the serum will appear positive only about 7 days after infection or later in severe coronavirus disease 2019 (COVID-19) [7,10]. It is difficult to detect early infections, which is not effective in blocking the source of early infection and increases the difficulty of preventing and controlling SARS-COV2 infection. In view of this situation, this study analyzed the positive rate of serum N protein before the generation of serum antibodies in patients with SARS-COV-2 infection, providing new diagnostic indicators for the early detection of SARS-COV-2 infection.

## 1. Materials and Methods

### 1.1 Specimen source

#### 1.1.1

50 samples of patients with pharyngeal swab or sputum SARS-COV-2 nucleic acid positive and serum SARS-COV-2 N protein antibody negative are collected from the First Affiliated Hospital of Anhui Medical University (20 cases) and Anhui Provincial Center for Disease Prevention and Control (30 cases), respectively. 37 samples of patients with pharyngeal swab or sputum SARS-COV-2 nucleic acid positive and serum SARS-COV-2 N protein antibody positive are from the First Affiliated Hospital of Anhui Medical University. Among the patients with negative serum N protein, the serum N protein antibody titer and CT results of 4 patients were analyzed.

#### 1.1.2

633 samples with pharyngeal swab or sputum SARS-COV-2 nucleic acid negative results and serum N protein antibody negative results are from the First Affiliated Hospital of Anhui Medical University, including 100 pregnant samples, 369 serum samples from other respiratory infections, 119 serum samples with increased rheumatoid factor and 45 hemolytic samples.

### 1.2. Reagents

#### 1.2.1

The SARS-COV-2 antigen quantitative detection kit (ELISA) (lot number 20200508) was developed by BIOHIT Healthcare (Hefei) Co., Ltd.

Principle: Double antibody sandwich method is used to detect SARS-CoV-2 N protein in human serum.

Results interpretation: The positive interpretation value (cutoff value) is 10.00 pg / mL. If the concentration of the specimen to be tested is less than 10.00 pg / mL, it is interpreted to be negative for SARS-COV-2 N protein. If the concentration of the specimen to be tested ≥ 10.00 pg / mL, it is interpreted as positive for SARS-COV-2 N protein.

#### 1.2.2

The SARS-COV-2 IgM / IgG antibody detection kit (colloidal gold method) (batch number SA200301) was developed by BIOHIT Healthcare (Hefei) Co., Ltd.

Principle: Colloidal gold immunochromatography method is used to detect anti-SARS-CoV-2 IgM / IgG N protein antibody in human serum.

Results interpretation: If the quality control line C developed color and the test line did not develop color, it is interpreted as negative. If the color of the quality control line C was developed, and the color of the test line was also developed, it is determined to be positive. If the quality control line C was not colored, the result is invalid and the sample needs to be re-tested.

### 1.3 Statistical analysis: ROC curve was drawn by SPSS 19.0

## 2. Results

### 2.1

The positive rate of serum N protein when pharyngeal swab or sputum nucleic acid results are positive and serum antibody results were negative for SARS-COV-2

50 cases of SARS-COV-2 nucleic acid positive and antibody negative patients had a serum N protein positive rate of 76%, including 2% with a concentration of 10.00-49.99 pg / mL, 8% with a concentration of 50.00-99.99 pg / mL, and 22% with a concentration of 100.00 – 299.99 pg / mL, and 44% with a concentrations 300.00 pg / mL. The negative rate of serum N protein is 24%, 12% had a concentration of 0.00-1.99 pg/mL, 10% had a concentration of 2.00-4.99 pg/mL, 2% had a concentration of 5.00-9.99 pg/mL. (See results in Table1)

**Table1.** Serum N protein composition ratio among Nucleic acid positive and antibody negative patients

| Detection marker | N Protein conc. | Percentage |
| --- | --- | --- |
| Nucleic acid positive + antibody | $\geq 10.00$ pg/mL | 76% ( 38/50 ) |
| Negative +N Protein positive | 10.00-49.99 pg/mL | 2% ( 1/50 ) |
|  | 50.00-99.99 pg/mL | 8% ( 4/50 ) |
|  | 100.00-299.99 pg/mL | 22% ( 11/50 ) |
|  | ≥300.00 pg/mL | 44% ( 22/50 ) |
| Nucleic acid positive + antibody | <10.00 pg/mL | 24% ( 12/50 ) |
| Negative +N Protein negative | 0.00-1.99 pg/mL | 12% ( 6/50 ) |
|  | 2.00-4.99 pg/mL | 10% ( 5/50 ) |
|  | 5.00-9.99 pg/mL | 2% ( 1/50 ) |

### 2.2

The positive rate of serum N protein when pharyngeal swab or sputum nucleic acid results are positive and serum antibody results were positive for SARS-COV-2

37 samples of patients with serum antibody positive after infection, they had a serum N protein positive rate of 2.7%, of which 2.7% had the concentration of 10.00-49.99 pg / mL and 0 % had the concentration of 50.00-99.99 pg / mL, 100.00-299.99 pg / mL, and ≥300.00 pg / mL.

The negative rate of N protein was 97.3 %, of which 73.0% had the concentration of 0.00-1.99pg / mL, 13.5 % had the concentration of 2.00-4.99pg / mL, and 10.8% had the concentration of 5.00-9.99pg / mL. (The results are shown in Table 2)

**Table2.**
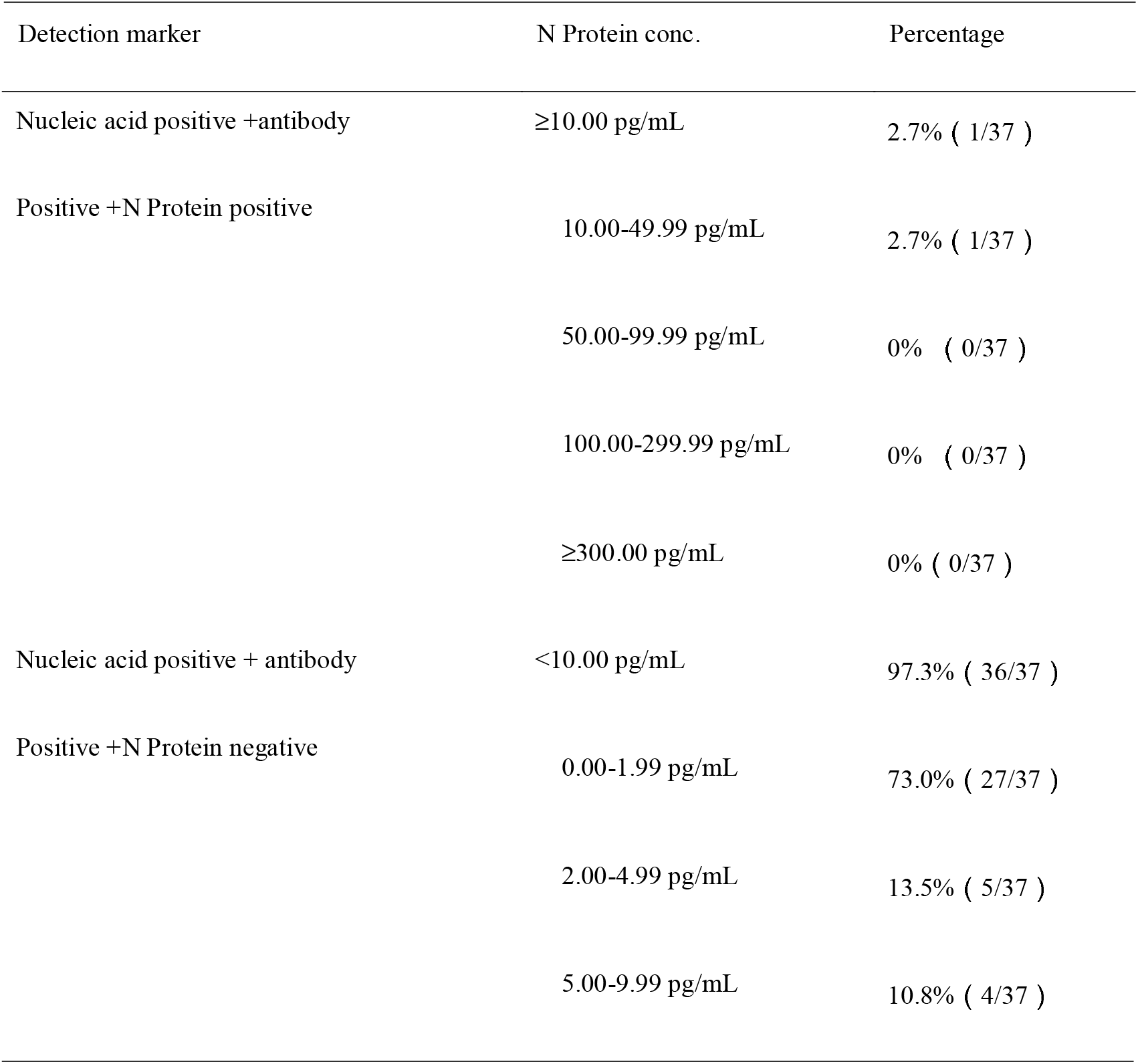
Serum N protein composition ratio among Nucleic acid positive and antibody positive patients

| Detection marker | N Protein conc. | Percentage |
| --- | --- | --- |
| Nucleic acid positive +antibody | ≥10.00 pg/mL | 2.7% ( 1/37 ) |
| Positive +N Protein positive | 10.00-49.99 pg/mL | 2.7% ( 1/37 ) |
|  | 50.00-99.99 pg/mL | 0% ( 0/37 ) |
|  | 100.00-299.99 pg/mL | 0% ( 0/37 ) |
|  | ≥300.00 pg/mL | 0% ( 0/37 ) |
| Nucleic acid positive + antibody | <10.00 pg/mL | 97.3% ( 36/37 ) |
| Positive +N Protein negative | 0.00-1.99 pg/mL | 73.0% ( 27/37 ) |
|  | 2.00-4.99 pg/mL | 13.5% ( 5/37 ) |
|  | 5.00-9.99 pg/mL | 10.8% ( 4/37 ) |

### 2.3 Analysis of antibody titer and CT results of 4 patients whose N protein is consistently negative

The serum antibody and CT results of 4 patients with consistent negative serum N protein results were tracked, the serum antibody was still negative or in low titer more than 9 days after the infected patients were admitted to the hospital. CT imaging showed that the inflammation of the lungs of the patients was mild or no obvious inflammation.

**Table3.** Antibody titer and CT results of 4 patients whose N protein is consistently negative

| Patient ID | Marker | 1-3 days after admission | 4-6 days after admission | 7-9 days after admission | >9 days after admission |
| --- | --- | --- | --- | --- | --- |
| 3 | Antibody titer | Negative | Negative | 1:1* | 1:1* |
|  | CT imaging | A little inflammation under the pleura of the right lower lobe and under pleura of the left upper lobe | A little inflammation under the pleura of the right lower lobe and in the upper left lung | Inflammatory lesions of left lung | A little inflammation of the left lung |
| 32 | Antibody titer | Negative | Negative | 1:1* | 1:1* |
|  | CT imaging | A little inflammation in the right upper lobe and left lower lobe | A little inflammation in the right upper lobe and left lower lobe, improved from previous exam | A little inflammation in the right upper lobe and left lower lobe, improved from previous exam | A little inflammation in the right upper lobe and left lower lobe, improved from previous exam |
| 33 | Antibody titer | 1:1* | 1:1* | 1:1* | 1:1* |
|  | CT imaging | No obvious lesions in both lungs | No obvious lesions in both lungs | No obvious lesions in both lungs | No obvious lesions in both lungs |
| 40 | Antibody titer | Negative | Negative | Negative | Negative |
|  | CT imaging | A little inflammation of the left upper lobe | A little inflammation of the left upper lobe, improved from the previous exam | A little inflammation of the left upper lobe, improved from the previous exam | A little inflammation of the left upper lobe, improved from the previous exam |

➤ 1:1 \*means when the serum is tested as undiluted, it showed weak positive results. Further dilution would result in negative reading.

### 2.4 Specificity of SARS-COV-2 serum N protein assay at the CUTOFF value recommended by the manufacturer

Serum N protein test results of 633 patients with non-SARS-COV-2 infection showed a specificity of 100%, including serum samples of 100 pregnant women, 369 samples with other respiratory infections, 119 serum samples with elevated rheumatoid factor, and 45 hemolytic samples. (The results are shown in Table 4)

**Table4.** Specificity of SARS-COV-2 serum N protein assay at the CUTOFF value recommended by

| Sample source | Number of antigen positive samples | No. of sample | Specificity |
| --- | --- | --- | --- |
| serum samples of pregnant women | 0 | 100 | 100% |
| serum samples with other respiratory infections | 0 | 369 | 100% |
| serum samples with elevated rheumatoid factor | 0 | 119 | 100% |
| hemolytic samples | 0 | 45 | 100% |
| Total | 0 | 633 | 100% |

### 2.5 Receiver operator characteristic curve (ROC curve)

Using SPSS 19.0 to calculate the receiver operating characteristic curve, the area under the curve was 0.9756 (95% confidence interval 0.9485 to 1.000, p <0.0001), sensitivity and specificity were 92% (95% confidence interval 81.16% to 96.85%) and 96.84% (95% confidence interval 95.17% to 97.15%). the best CUTOFF value is 1.850 pg / mL. (The results are shown in Figure 1)

**Figure 1.**
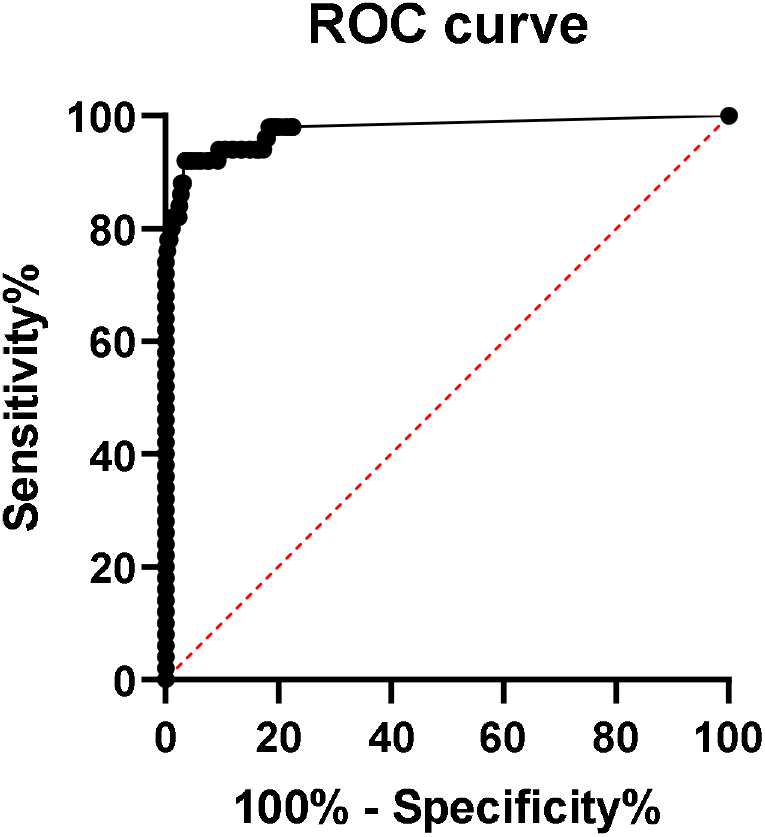
ROC curve of serum SARS-COV-2 N protein, area under the ROC curve is 0.9756, 95% confidence interval (CI) is 0.9485 to 1.000, p value is lower 0.0001, sensitivity and specificity are 92% (95% CI 81.16% to 96.85%) and 96.84% (95% CI 95.17% to 97.15%), respectively. The best CUTOFF value is 1.850 pg/mL.

When calculating the CUTOFF value using the ROC curve, the specificity of pregnancy population samples, other respiratory infection samples, elevated rheumatoid factor samples, and hemolytic samples were 91.0%, 97.3%, 100%, and 97.8%, respectively. (The results are shown in Table 5)

**Table5.** The specificity of SARS-COV-2 serum N protein assay when CUTOFF value is calculated by using the ROC curve

| Sample type | No. of sample | No. of N protein positive sample | Specificity |
| --- | --- | --- | --- |
| serum samples of pregnant women | 100 | 9 | 91.0% |
| serum samples with other respiratory infections | 369 | 10 | 97.3% |
| serum samples with elevated rheumatoid factor | 119 | 0 | 100% |
| hemolytic samples | 45 | 1 | 97.8% |
| Total | 633 | 20 | 96.8% |

### 2.6 Distribution of N Protein concentration result of sample data

Figure 2 shows the distribution of sample results using the CUTOFF value equaling to 10.00 pg/mL. or 1.850 pg/mL. Group A (Nucleic Acid Positive + Antibody Negative, N=50) shows the distribution of samples is mostly above 10.00 pg/mL, with a large spread from zero. Group B (Nucleic Acid Positive + Antibody Positive, N=37) and Control Group (non-SARS-COV-2 infection, N = 633) shows the distribution of samples is mostly below 10.00 pg/mL, with a cluster of samples around zero.

**Figure 2.**
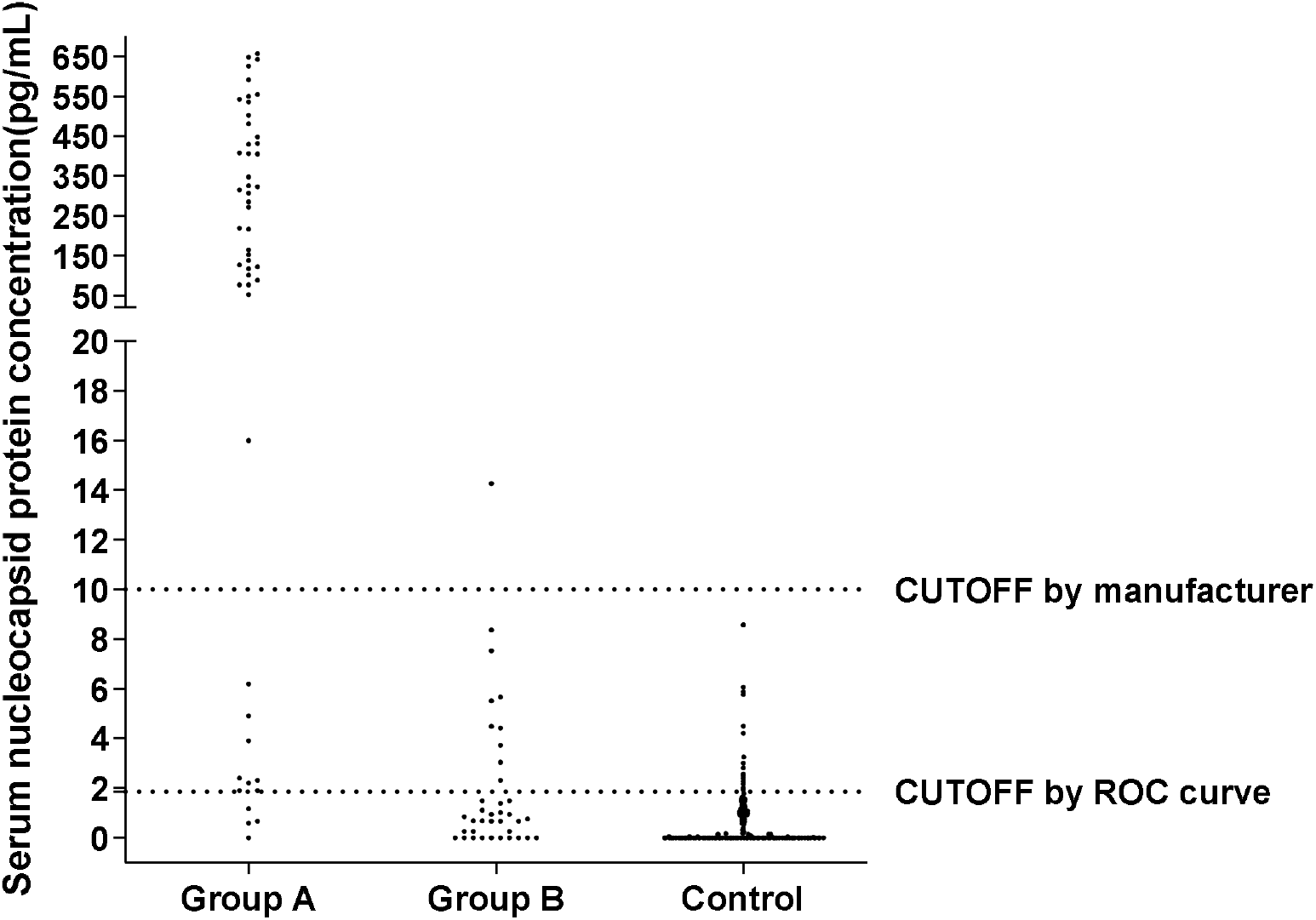
Distribution of N Protein concentration result of sample data using the CUTOFF value equaling to 10, Group A (Nucleic Acid Positive + Antibody Negative, N=50), Group B (Nucleic Acid Positive + Antibody Positive, N=37), Control Group (non-SARS-COV-2 infection, N = 633)

## 3. Discussion

Pulmonary inflammation is most common among SARS-CoV-2 infection, and it can also invade other tissues [11,12]. Some patients have viremia. It was reported that SARS-CoV-2 RNA was detected in the blood among 15% of the 41 COVID-19 patients who were admitted to hospital for the first time in Wuhan [13]. The results of this study showed that 76.8% of patients diagnosed with SARS-CoV-2 infection were positive for N protein before the emergence of the N antibody (see Table 1). Further studies are needed to confirm whether the infected person has a higher incidence of viremia in the early stage or whether the over-expressed N protein of the lung virus spills into the blood.Serum SARS-CoV-2 antibody test has been widely recognized as serological evidence for the diagnosis of COVID-19 [14-16], but due to the lag in the development of antibodies, the clinical diagnosis “window period” is too long, and serum SARS-COV-2 antibody test is not suitable for the early diagnosis of SARS-CoV-2 infection [10,15]. Our research results show that when the serum antibody is positive, the serum N protein positive rate is only 2.7%, and the overall serum N protein concentration value is low (see Table 2), suggesting that the detection of serum antibody and N protein has good complementary effects for the diagnosis of SARS-CoV-2 infection. The results in Table 3 show that 4 early serum N protein-negative patients had low antibody titers and CT showed that the lesions were mild or no obvious inflammation. Whether it is related to viral load in vivo remains to be further studied.

The results in Table 1, Table 4 and Table 5 showed that using the CUTOFF value recommended by the manufacturer, the specificity of serum N protein assay can reach 100%, but the sensitivity is only 76.8%. If the ROC curve is used to establish a personalized CUTOFF value, its specificity decreases to 96.84%, but its sensitivity increases to 92%. Therefore, we suggest that laboratories should select appropriate CUTOFF values according to the intended use. If you want to achieve higher diagnostic specificity, we recommend the use of CUTOFF values recommended by manufacturer. If you want to find as many infected people as possible and establish the basis for controlling the source of infection. It is recommended to select the CUTOFF value obtained from the ROC curve.

The results of this study provide a laboratory basis for serum SARS-CoV-2 N protein detection for early diagnosis of infection. Compared with SARS-CoV-2 RNA detection from pharyngeal swab or sputum, serum SARS-CoV-2 N protein detection has obvious advantages in specimens collection and treatment. For example, N protein has better stability than RNA, which may effectively make up for missed diagnosis caused by RNA false negative results due to various reasons. Although the number of cases in this study was small, our results are extremely encouraging, which brings a new thought to the current incomplete diagnosis of SARS-CoV-2 infection. Combined detection of three markers: pharyngeal swab or sputum SARS-CoV-2 RNA, serum N protein and serum antibody may be the direction of diagnosing SARS-CoV-2 infection in the future.

## 4. Conclusion

### 4.1

The detection of SARS-COV-2 serum N protein has a high diagnostic value for infected patients before the appearance of antibodies, and shortens the window of serological diagnosis.

### 4.2

According to the CUTOFF value recommended by the manufacturer, the specificity of SARS-COV-2 serum N protein detection is 100%, and the sensitivity is 76.8% before the antibody appearance.

### 4.3

Determining the CUTOFF value according to the ROC curve. The specificity of the SARS-COV-2 serum N protein detection was 96.84%, and the sensitivity was 92% before the antibody appearance.

### 4.4

It is recommended that each laboratory establishes its own CUTOFF value according to the purpose of the application.

## Data Availability

The data that support the findings of this study are available from the corresponding author on reasonable request. Participant data without names and identifiers will be made available after approval from the corresponding author and National Health Commission. After publication of study findings, the data will be available for others to request. The research team will provide an email address for
communication once the data are approved to be shared with others. The proposal with detailed description of study objectives and statistical analysis plan will be needed for evaluation of the reasonability to request for our data. The corresponding author and National Health Commission will make a decision based on these materials. Additional materials may also be required during the process.

## Contributors

TL and LW had the idea for and designed the study and had full access to all data in the study and take responsibility for the integrity of the data and the accuracy of the data analysis. HW, XL and SZ contributed to writing of the report. YX and WW contributed to the statistical analysis. All authors contributed to data acquisition, data analysis, or data interpretation, and reviewed and approved the final version.

## Disclaimer

The antibody screening, methodological selection and reagent production of this study were completed in the research and development department of Biohit Healthcare (Hefei) Co., Ltd. Methodological evaluation and performance verification were completed in the First Affiliated Hospital of Anhui Medical University. All authors have no conflicts of interest.

## Ethics

This study was approved by the Ethics Committee of the First Affiliated Hospital of Anhui Medical University.

## Fund

Supported by the Scientific Research Project of Anhui Province for the Prevention and Control of New Coronavirus Pneumonia (202004a07020015).

